# Applications of Artificial Intelligence in Vasculitides: A Systematic Review

**DOI:** 10.1101/2024.10.07.24314995

**Authors:** Mahmud Omar, Reem Agbareia, Mohammad E. Naffaa, Abdulla Watad, Benjamin S Glicksberg, Girish N Nadkarni, Eyal Klang

## Abstract

**Background and Aim:** Vasculitides are rare inflammatory disorders that sometimes can be difficult to diagnose due to their diverse presentations. This review examines the use of Artificial Intelligence (AI) to improve diagnosis and outcome prediction in vasculitis.

**Methods:** A systematic search of PubMed, Embase, Web of Science, IEEE Xplore, and Scopus identified relevant studies from 2000 to 2024. AI applications were categorized by data type (clinical, imaging, textual) and by task (diagnosis or prediction). Studies were assessed for risk of bias using PROBAST and QUADAS-2 tools.

**Results:** Forty-six studies were included. AI models achieved high diagnostic performance in Kawasaki Disease, with sensitivities up to 92.5% and specificities up to 97.3%. Predictive models for complications, such as IVIG resistance in Kawasaki Disease, showed AUCs between 0.716 and 0.834. Other vasculitis types, especially those using imaging data, were less studied and often limited by small datasets.

**Conclusion:** The current literature shows that AI algorithms can enhance vasculitis diagnosis and prediction, with deep and machine learning models showing promise in Kawasaki Disease. However, broader datasets, more external validation, and the integration of newer models like LLMs are needed to advance their clinical applicability across different vasculitis types.

## Introduction

Vasculitides are systemic inflammatory and immune-mediated disorders affecting blood vessels that pose many diagnostic challenges due to their varied presentation patterns (1,2). Early diagnosis and treatment are critical for improving patient outcomes (3). Artificial intelligence (AI) has emerged as a potential tool to address these challenges in vasculitis care.

AI techniques—such as machine learning, deep learning, and natural language processing (NLP)—can analyze medical data to assist in diagnosis, predict outcomes, and personalize treatment plans (4,5). These methods can process information from clinical records, laboratory results, imaging studies, and beyond (6). By identifying subtle patterns and associations from patients, AI may expedite vasculitis diagnosis, enabling earlier interventions (7). Although some AI models have shown promise in related fields, such as predicting the need for autoantibody testing in systemic autoimmune diseases (6–9) or detecting anti-neutrophil cytoplasmic antibodies positivity from free-written text with accuracy comparable to human analysis (10), a gap remains in the application of AI to rare diseases like vasculitides. The rarity of these conditions makes them less familiar to clinicians, leading to higher rates of missed or delayed diagnoses (11). This gap underscores the need for AI models specifically designed to enhance the diagnosis, monitoring, and treatment of vasculitis. This systematic literature review aims to address this gap by mapping current AI applications in vasculitis care and offering future directions.

## AI Foundational Concepts

### AI Algorithms and Data Types

This review focuses on two main categories of AI algorithms—*machine learning (ML)* and *deep learning (DL)*. These algorithms are applied to textual data through *natural language processing (NLP)*, imaging data using *computer vision*, and *clinical and bioinformatical tabular data* such as laboratory results, patient records, and genetic data. Together, these techniques support diagnosis, prognosis, and treatment personalization in vasculitis care (**Figure 1**).

**Figure 1:**
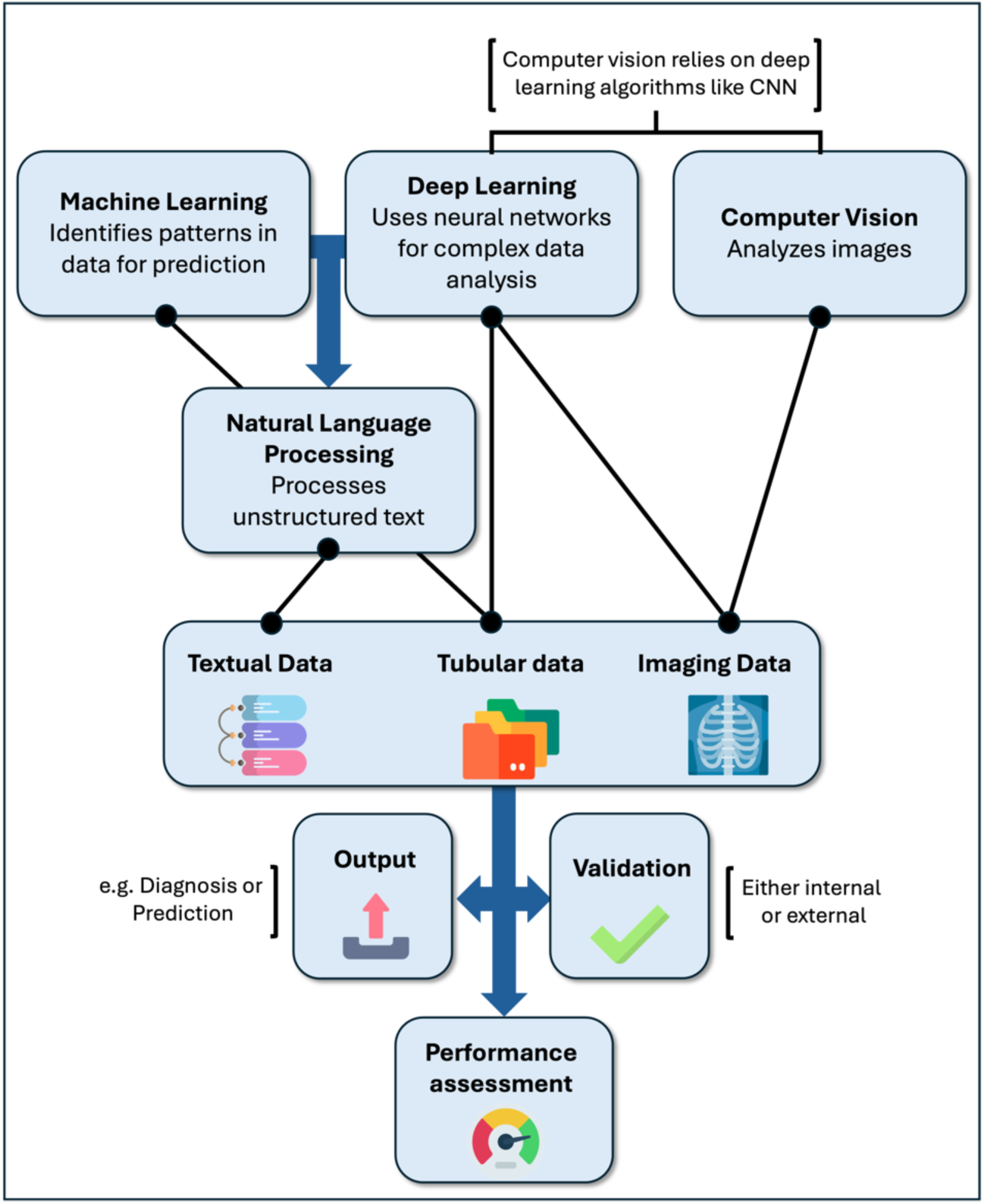
Flowchart representing the different AI models and datatypes.

### Machine Learning (ML)

ML models find patterns in data to make predictions or decisions (12). Linear algorithms include linear and logistic regression, and Support Vector Machines (SVMs). These methods look for the best lines or boundaries (called hyperplanes) to separate data into different groups. There are types of SVMs that can handle both simple (linear) and more complex (non-linear) problems (12).

Many of today’s reliable ML models are *decision trees* based. A tree-based model, like a decision tree, asks a series of yes/no questions about the data. Each question splits the data into smaller parts, forming branches like a tree (12). *Random Forest* is a common tree-based model. It creates many decision trees and combines their results to make better predictions. *Random Forest* works well with many kinds of data and is less likely to overfit, meaning it doesn’t get too attuned by training data to underperform on the testing data (12). *Gradient Boosting models*, like XGBoost and LightGBM, work differently. They build a series of weak models, typically decision trees, one after the other. Each new model tries to fix the mistakes made by the previous one. This step-by-step process helps make the final model more accurate and efficient (13).

### Computer vision

Computer vision is a field of AI that enables machines to interpret and understand visual data from the world, such as images or videos (14). It involves techniques for recognizing objects, identifying patterns, and analyzing scenes, allowing computers to make decisions or perform actions based on visual input. Applications include facial recognition, autonomous vehicles, medical imaging, and more. It relies on deep learning models, particularly convolutional neural networks (CNNs), to process and analyze visual information efficiently (14).

### Deep Learning (DL)

DL models use *artificial neural networks (ANNs)*, which are inspired by the way the brain works. These networks have many layers, allowing them to find complex patterns in data (15). *Convolutional Neural Networks* (CNNs) are a special type of DL model used mainly in computer vision for image data. They automatically learn important features from images by applying filters, which helps the model focus on key details like edges or shapes (16). *U-Net* is a type of CNN that is specifically designed for tasks like biomedical image segmentation. The “U” in U-Net refers to its unique structure: it has two main parts. In the first part, the network breaks the image down layer by layer, learning more abstract features as it goes. In the second part, it “rebuilds” the image layer by layer, focusing on detailed localization. This structure looks like the letter “U” when visualized, and it helps the model precisely segment the boundaries of objects, like organs or tumors, in medical images (17). *Vision Transformers* are another DL model used for images. They adapt a technique originally used for text data and apply it to image analysis (17).

### Natural Language Processing (NLP)

NLP uses both traditional ML models and more advanced DL models to allow computers to understand and work with human language (18). *ML-based* algorithms like logistic regression, which are used for simpler tasks like spam detection or document classification. DL-based models include Recurrent Neural Networks (RNNs) and Transformers, which handle more complex tasks such as machine translation or text summarization (19). One of the key advancements in NLP is the *attention mechanism*. Attention allows the model to focus on the most important parts of the input data. For example, when translating a sentence, the attention mechanism helps the model focus on the most relevant words in the sentence, even if they are far apart. This helps the model better understand the context and meaning behind each word (19).

*Transformers* are built around this attention mechanism. Unlike older models, Transformers can process an entire sequence (like a sentence or paragraph) all at once. They don’t just look at words in order; instead, they focus on relationships between words, making them much better at understanding context. This is why Transformers are widely used in tasks like language translation, question-answering, and text generation (19). Building on Transformers, *Large Language Models (LLMs)* like *Generative pretrained transformer (GPT)* (used in *ChatGPT*) and *Bidirectional Encoder Representations from Transformers (BERT)* are pre-trained on vast amounts of text data (20). These models can perform many NLP tasks such as generating text, summarizing documents, or answering complex questions. LLMs use the attention mechanism to understand large and complex texts in a way that mimics human comprehension, making them extremely versatile across a range of language-based tasks (20).

### Model Performance and Validation

ML and DL models in medical studies are measured using different metrics to see how well they work. *Area Under the Curve (AUC)* shows how good the model is in discriminating two groups apart, like sick versus healthy patients. The closer the AUC is to 1, the better the model (21). *Sensitivity (Recall)* measures how well the model finds all the cases with the condition (21). *Specificity* shows how well a model identifies cases without the condition (21). *Accuracy* is the percentage of correct predictions overall (22). *Positive Predictive Value (PPV)* shows how many of the predicted positives are actually positive (21). *Negative Predictive Value (NPV)* shows how many of the predicted negatives are actually negative (21). *F1-score* balances precision and recall, making it valuable when there’s an imbalance between classes, such as in rare diseases (21). *Dice coefficient* is used in image analysis to check how well the model has segmented an image, comparing how similar the predicted segmentation is to the true segmentation (23).

To make sure the model is reliable, researchers use different validation methods. *Internal validation* includes methods like k-fold cross-validation. In this method, the data is split into k groups, and the model is trained and tested multiple times, using a different group each time for testing (23). *External validation* tests the model on data from other hospitals or institutions to see how well it works with new, unseen data (23).

## Materials and Methods

### Registration and Protocol

We conducted a systematic review following the Preferred Reporting Items for Systematic Reviews and Meta-Analyses (PRISMA) guidelines (24). The protocol was registered with the International Prospective Register of Systematic Reviews (PROSPERO), registration number: CRD42024591969 (25).

### Data Sources and Search Strategy

We searched PubMed, Embase, Web of Science, IEEE Xplore, and Scopus for studies published between January 1, 2000, and September 18, 2024. Our search combined AI-related terms with vasculitis terms. We supplemented database searches with manual screening of reference lists from included studies. The full search strategy is available in the **Supplemental Materials**. We developed our search strategy following the methods outlined in Chapter 4 of the Cochrane Handbook for Systematic Reviews of Interventions (version 6.4) (26). We used the Rayyan web application for initial screening (27).

### Study Selection

Two reviewers (MO and EK) independently screened titles and abstracts using Rayyan. We obtained full texts for potentially eligible studies. The same reviewers then assessed these for inclusion. We resolved disagreements through discussion or third-reviewer arbitration. We included peer-reviewed studies that evaluated any direct AI applications in patients with any type of Vasculitides. We excluded non-peer-reviewed materials.

### Data Extraction and Quality Assessment

We developed a standardized form for data extraction. One reviewer extracted data, which was verified by a second reviewer. We extracted information on study design, AI category and model, vasculitis type, measurement methods, type and source of data, validation methods and results, and key findings. Two tools were used to assess the risk of bias in the included studies. PROBAST was applied to evaluate prediction model studies, while QUADAS-2 was used for diagnostic accuracy studies. PROBAST assesses four domains: participants, predictors, outcome, and analysis (28). QUADAS-2 evaluates three domains: patient selection, index test, and reference standard (29). The flow and timing domain of QUADAS-2 was not assessed as it is less relevant for AI studies in this context. Each domain was rated as low, high, or unclear risk of bias.

### Data analysis

Meta-analysis was not feasible due to the heterogeneity in study designs, populations, AI methods, and outcomes across the included studies. Therefore, a narrative synthesis was conducted. We grouped studies by AI application type (diagnosis or prediction) and data type (clinical and bioinformatical data, imaging, or unstructured textual data). Within each group, key study characteristics, AI models, performance metrics, and validation methods were summarized and compared.

## Results

### Search Results and Study Selection

A total of 681 studies were identified across five databases during the initial screening. After removing 137 duplicates and excluding 477 studies based on title and abstract screening, 67 studies were selected for full-text retrieval. Of these, 46 studies were retrieved and included in the systematic review. **Figure 2** shows the PRISMA flowchart.

**Figure 2:**
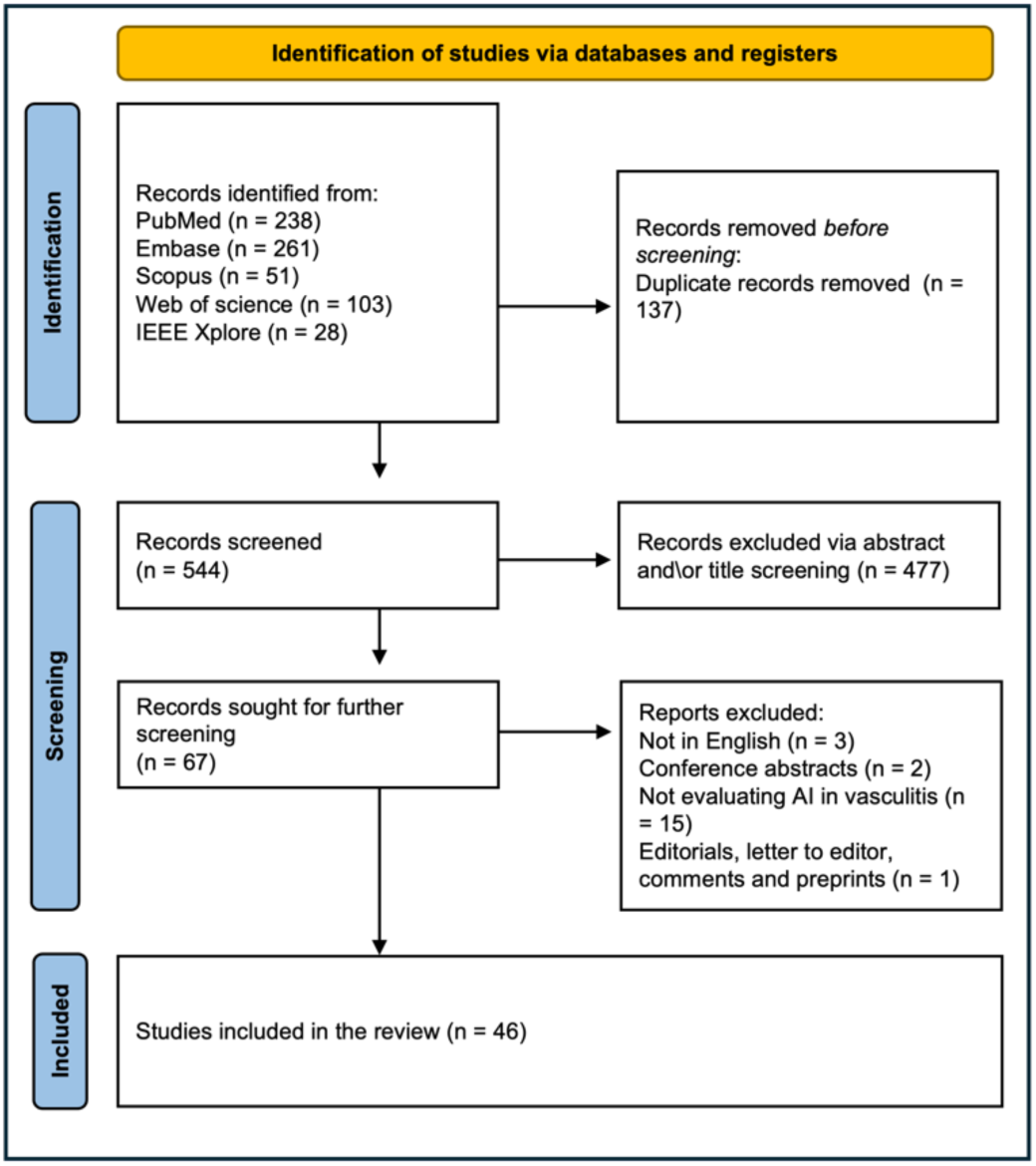
PRISMA flowchart.

### Overview of the included studies

This review includes 46 studies (30–75), published between 1994 and 2024 that investigate the application of AI in vasculitis. We classified these studies into two main categories: diagnosis and prediction. Diagnosis includes any AI application for diagnosing, classifying, or differentiating vasculitis, encompassing 28 studies. Prediction involves any AI application predicting complications or outcomes in vasculitis patients, totaling 18 studies (**Tables 1 and 2**). The studies are further grouped based on the data types used: structured clinical and bioinformatical data (including genetic data) (31 studies), imaging data, analyzed using computer vision techniques, mainly DL (12 studies), and clinical texts, which are analyzed using NLP techniques (3 studies).

Sample sizes in these studies ranged from small cohorts of 6 patients to large datasets exceeding 70,000 individuals. The ML models employed included Random Forest, SVM, Extreme Gradient Boosting (XGBoost), and Light Gradient Boosting Machine (LightGBM). DL models included CNNs, U-Net architectures, and custom neural networks, among others. Data sources varied widely, encompassing clinical data, imaging data (such as echocardiograms, fluorescein angiography, optical coherence tomography), laboratory results, gene expression profiles, and electronic health records (**Figure 3**).

**Figure 3:**
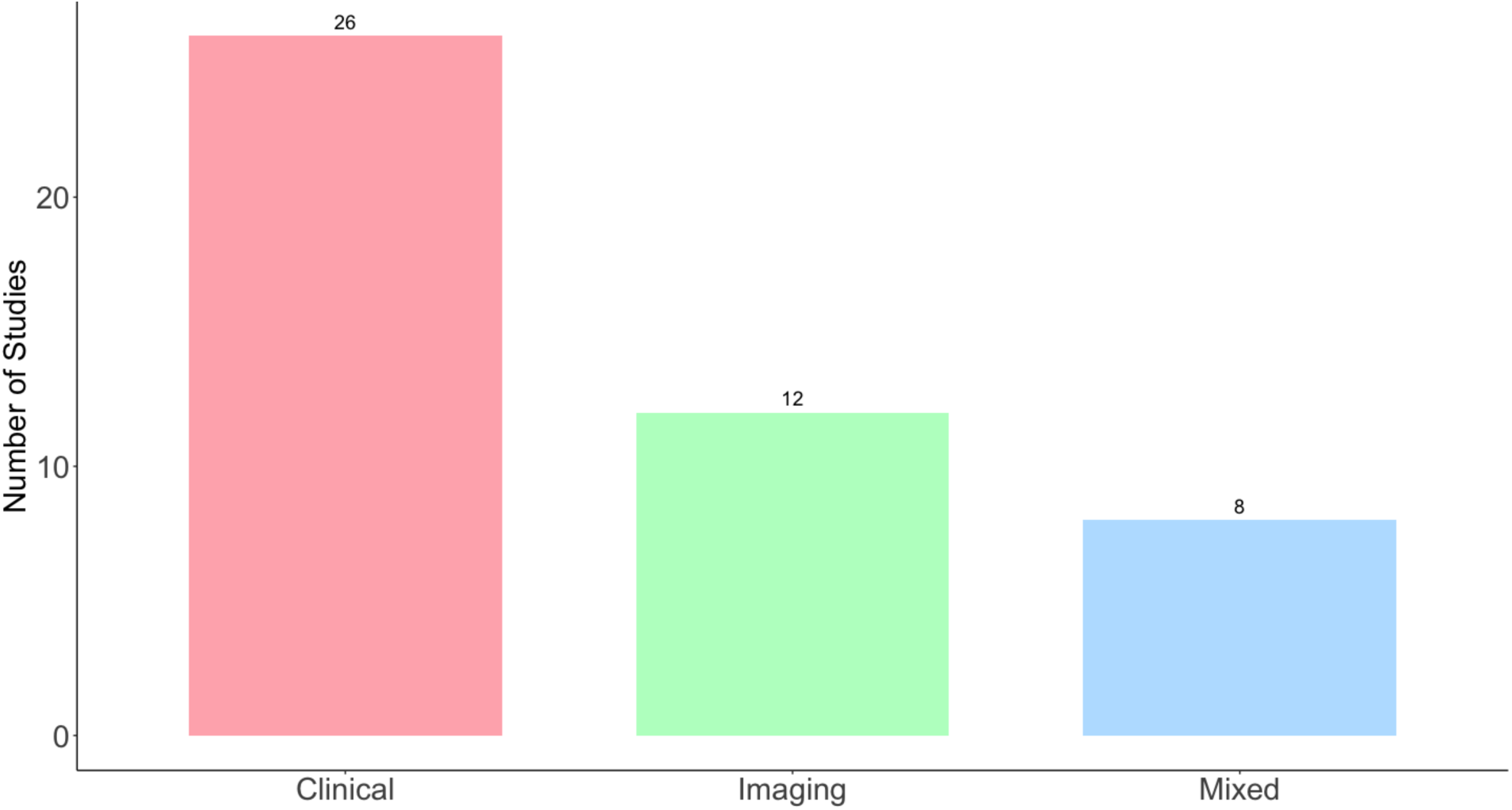
Frequencies of the different data types utilized in the included studies.

As expected, a surge of AI publications can be seen in 2024, as shown in Figure 4.

**Figure 4:**
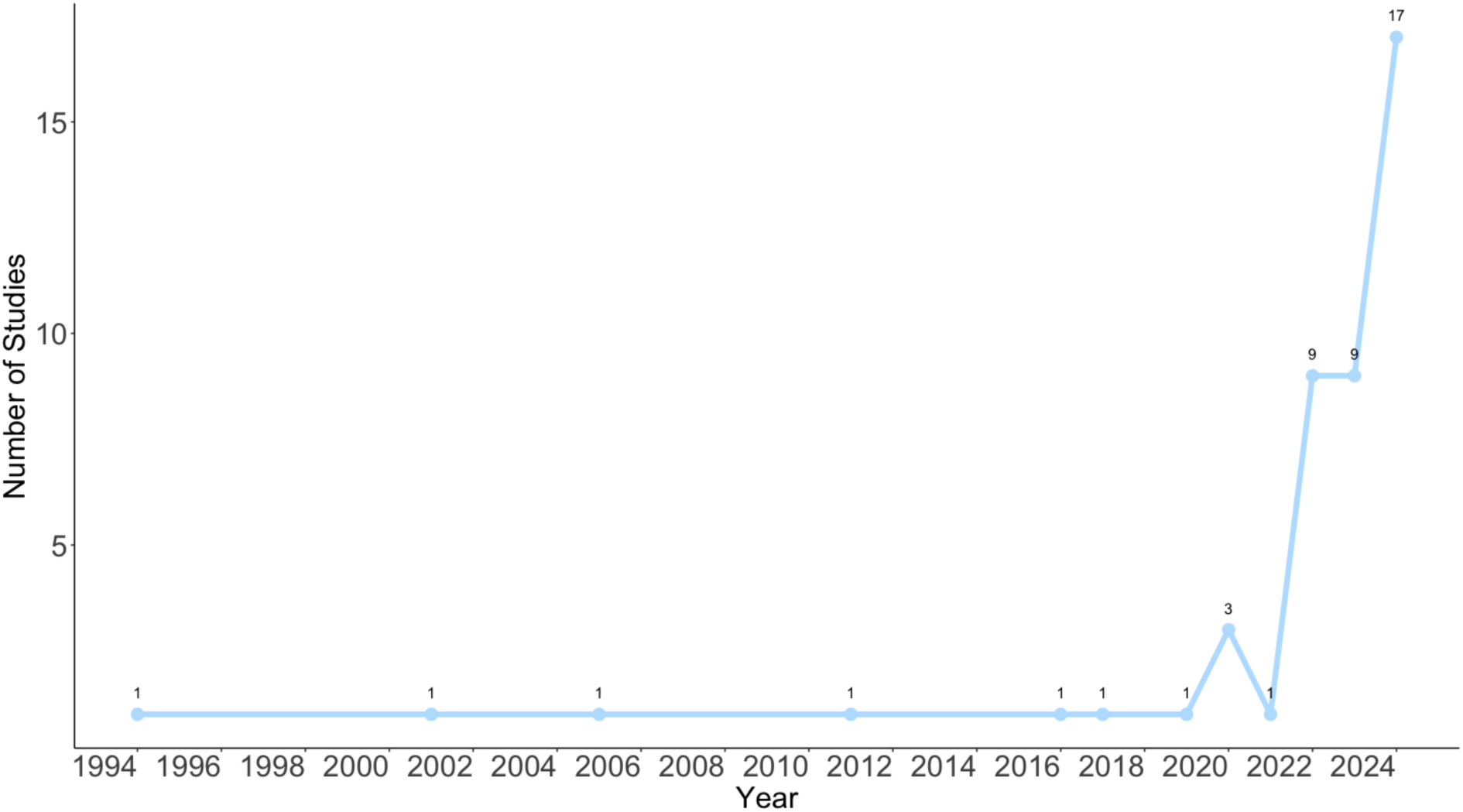
Number of published studies over time.

**Tables S3** in the supplement offers a detailed summary of the included studies.

### Risk of bias assessment

**Table S1** summarizes the PROBAST assessment, where a total of 19 studies were evaluated. Of these, 10 studies (52.6%) were rated as having a low risk of bias, 9 studies (47.4%) were judged to have a high risk of bias, and none were classified as unclear. For the diagnostic accuracy studies, QUADAS-2 was applied, as summarized in **Table S2**. Out of 27 studies, 13 (48.1%) had a low risk of bias, while 14 (51.9%) were rated as having a high risk of bias, with no studies marked as unclear. Across all assessments, there were 124 instances of low risk (79.5%), 31 instances of high risk (19.9%), and 2 instances of unclear risk (1.3%).

## Clinical and Bioinformatical Data

### Diagnosis

Thirteen studies applied machine learning techniques for the diagnosis of Vasculitis.

### Kawasaki Disease Diagnosis

Four studies focused on Kawasaki Disease (KD). Tsai et al. conducted a large-scale study using XGBoost to differentiate KD from other febrile conditions in 74,641 children. Their model achieved 92.5% sensitivity and 97.3% specificity using laboratory parameters such as pyuria, white blood cell count in urine, Alanine Transaminase (ALT) levels, and C-reactive protein (CRP) levels (72). Similarly, Chi Li et al. distinguished KD from sepsis in 608 patients using least absolute shrinkage and selection operator (LASSO) and SVM techniques. Their model achieved an AUC of 0.878 in the validation set, utilizing similar clinical and biochemical parameters. The study included 299 KD patients and 309 sepsis patients (33). Additionally, Duan et al. developed an Explainable Boosting Machine to diagnose KD in 4,087 pediatric patients, including 2,971 with KD and 1,116 febrile children. The model achieved 93% accuracy using clinical and laboratory data (62).

Using a different data type, Zhang et al. combined Random Forest and neural networks to diagnose KD using gene expression data from multiple public datasets. They reported perfect classification in KD patients. The study analyzed data from five Gene Expression Omnibus datasets, including 78 KD patients and 55 healthy controls in one dataset, and 76 KD patients and 37 healthy controls in another (53).

### Other Vasculitides

Astion et al. used a neural network to classify Giant Cell Arteritis (GCA) and other vasculitis types in 807 patients (214 GCA cases and 593 other vasculitis cases). The network correctly classified 94.4% of GCA cases using eight clinical criteria, including age, localized headache, temporal artery tenderness, and erythrocyte sedimentation rate (ESR) (38). In another study focusing on GCA, Vries et al. used radiomics features from FDG-PET scans to differentiate GCA from atherosclerosis. They analyzed 20 [18F]FDG-PET scans from 20 patients (10 with GCA and 10 with atherosclerosis), segmented into 80 aortic segments. The model achieved 90% accuracy (54).

Schmitt et al. applied a neural network to distinguish between Eosinophilic granulomatosis with polyangiitis (EGPA) and Granulomatosis with polyangiitis (GPA) in 80 patients (40 with EGPA and 40 with GPA). Using 43 clinical and laboratory parameters, they achieved 96% accuracy during validation (61). Using similar methods, Linder et al. differentiated GPA from microscopic polyangiitis (MPA) in 385 patients using various ML methods. Their neural network model surpassed 94% accuracy using 23 clinical measurements and ANCA serology. The study included patients from both a multicenter cohort (240 GPA, 78 MPA) and a monocenter cohort (46 GPA, 21 MPA) (55).

Regarding Behçet’s disease, Güler et al. analyzed ophthalmic arterial Doppler signals in 106 subjects (54 with Behçet’s disease and 52 healthy controls) to detect Behçet’s disease. Their multilayer perceptron neural network achieved 96.43% accuracy for healthy subjects and 93.75% accuracy for patients with ocular Behçet’s disease (49).

Nie et al. used XGBoost to differentiate abdominal Henoch-Schönlein Purpura (AHSP) from acute appendicitis in 6,965 children (2,201 with AHSP and 4,764 with acute appendicitis). The model achieved 82.4% accuracy using 53 laboratory indicators (39).

Regarding studies focusing on wider range of Vasculitides, Morris et al. combined Raman spectroscopy with ML algorithms to detect histological lesions in ANCA-associated glomerulonephritis. They analyzed 27 renal biopsy samples and 10 paired urine samples from 28 patients. The model showed high accuracy for tissue samples (sensitivity and specificity up to 100% for some lesions) but limited performance for urine samples (40).

Ryyppö et al. employed ML models to identify vasculitis, myositis, and glomerulonephritis in 114,897 patients, including 2,919 with vasculitides, and 100,000 controls. Their XGBoost model achieved a true positive rate of 76.7% and a true negative rate of 98.4% for vasculitis detection using clinical and laboratory data (43).

### Prediction

Eighteen studies utilized ML for prediction tasks in vasculitis, focusing on forecasting treatment resistance, organ involvement, disease activity, and complications.

### Predicting IVIG Resistance in Kawasaki Disease

Seven studies concentrated on predicting resistance to intravenous immunoglobulin (IVIG) in KD patients. Wang et al. (2024) analyzed 1,271 patients using Random Forest. They used laboratory indicators including CRP, and procalcitonin, achieving an AUC of 0.816 in the training set and 0.800 in the validation set (51). Similarly, Sunaga et al. created a scoring system based on LightGBM, achieving an AUC of 0.78. Their model incorporated demographic variables, laboratory data, and echocardiographic parameters (36).

Miyagi et al. studied 1,225 patients, including 228 IVIG non-responders. They developed a stacking classifier with an AUC of 0.716, identifying influential predictors such as sodium level and neutrophil-to-lymphocyte ratio (67).

Kuniyoshi et al. focused on a smaller cohort of 98 children, with 20 resistant to IVIG. They compared Logistic Regression, SVM, and XGBoost using demographic details and laboratory values, with AUCs ranging from 0.58 to 0.75 (44). Deng et al. examined 602 patients (68 IVIG-resistant) using XGBoost. They achieved an AUC of 0.82, and identified sodium levels and hemoglobin as significant predictors (45).

Joung et al. created a decision tree model using data from 896 KD patients. They predicted IVIG resistance and coronary artery dilatation using clinical and laboratory data, including total bilirubin and N-Terminal pro-Brain Natriuretic Peptide (NT-proBNP) levels. Their model showed an evaluation accuracy of 90.5% for IVIG resistance and 90.3% for coronary artery dilatation. The ROC-AUC for IVIG resistance was 0.834 (95% CI: 0.675–0.973) (30).

### Predicting Complications and Disease Flare-ups

Cao et al. used XGBoost to predict renal damage in 240 children with Henoch-Schönlein Purpura. They incorporated clinical data such as patient symptoms, age, recurrence of purpura, and laboratory data. Their model achieved an AUC of 0.895 in the training set and 0.870 in the test set (46). Similarly, Wang et al. (2022) applied Random Forest to predict renal damage in 288 children with IgA vasculitis. They used 42 indicators including demographic characteristics, clinical symptoms, and laboratory tests. The Random Forest model demonstrated the highest predictive performance with an accuracy of 0.83, recall of 0.86, and AUC of 0.91 (69). In another study focusing on predicting renal damage in IgA vasculitis, Pan et al developed a Random Forest, analyzing 217 inpatients for training and 46 for validation. Their model achieved an accuracy of 91%, and an AUC-ROC of 0.94 (65).

Lu et al. predicted severe ischemic complications in 703 Takayasu arteritis patients, of which 97 (13.8%) experienced complications such as stroke and myocardial infarction. They used clinical data including age, Body mass index (BMI), cardiovascular risk factors, blood pressure, and renal artery involvement. The best-performing model, Bagged FDA using gCV Pruning, achieved an AUC of 0.773 in the validation cohort. A Weighted Subspace Random Forest model also performed well, with an AUC of 0.770 (50). On another study regarding the prediction of coronary involvement, Azuma et al. analyzed 314 KD patients to predict coronary artery lesions. Their neural network used similar clinical data. The model achieved a sensitivity of 73%, specificity of 99% for predicting coronary artery lesions (71). Another similar study was carried out by Tang et al. They examined 158 children with KD using Random Forest, incorporating demographic characteristics, signs, symptoms, and laboratory results. Their model achieved an AUC of 0.925 and an accuracy of 0.930 (95% CI, 0.905 to 0.956) in predicting coronary artery lesions. They compared this to logistic regression (AUC 0.888) and XGBoost (AUC 0.879) models (68).

Ing et al. analyzed 1,833 patients who underwent temporal artery biopsy to predict biopsy-proven GCA. They used clinical data including age, gender, headache, temporal artery abnormality, and laboratory results. Their neural network model achieved an AUC of 0.860, sensitivity of 69.5%, and specificity of 89.1%. Comparatively, their logistic regression model achieved an AUC of 0.867, sensitivity of 52.5%, and specificity of 95.1% (63). In another study focusing on GCA, Venerito et al. studied 107 GCA patients to predict disease flare after glucocorticoid tapering. They used clinical data from electronic health records and laboratory data such as ESR and CRP levels. Their Random Forest model showed the best performance with 71.4% accuracy and an AUC of 0.76. The model demonstrated higher precision compared to logistic regression (62.6%) and decision tree (50.8%) models (75).

Hammam et al. (2023) developed an XGBoost model to predict vision-threatening Behçet’s disease in 1,094 patients, with 549 (50.2%) having vision-threatening complications. They used clinical and demographic variables to identify patients at risk. The XGBoost model achieved an AUC of 0.85 (95% CI: 0.81–0.90), accuracy of 85%, sensitivity of 85%, and specificity of 86% (52).

## Computer Vision

Twelve studies employed DL techniques for the diagnosis of vasculitis, utilizing CNNs and other DL architectures to analyze imaging data.

### Kawasaki Disease Imaging

Abdolmanafi et al. (2017) analyzed optical coherence tomography (OCT) images from 26 patients with KD, extracting 4,800 regions of interest. They used pre-trained CNNs as feature extractors combined with Random Forest classifiers to classify coronary artery layers (48). The model achieved 96% accuracy in detecting the media layer. In a follow-up study, Abdolmanafi et al. (2020) expanded their approach, analyzing 5,040 OCT frames from 45 pullbacks. They employed CNNs and fully convolutional networks for automatic diagnosis of coronary artery lesions in KD, achieving 90% accuracy for lesion characterization (37).

Rodríguez et al. focused on automatic segmentation of coronary arteries in echocardiograms for KD diagnosis. They analyzed 1,531 images from 36 anonymized Digital Imaging and Communications in Medicine (DICOM) files, using various U-net architectures. Their best-performing model, ResNet50-U-net with transfer learning, achieved 99.76% accuracy, outperforming traditional manual segmentation methods (47).

Lee et al. (2024) employed the Multiple Receptive Attention Network (*MRANet*) to differentiate incomplete KD from pneumonia. They analyzed 147 echocardiographic images (88 KD cases, 59 pneumonia cases), achieving 78.82% accuracy. The MRANet showed comparable sensitivity to experienced cardiologists (81.82% vs. 85%) and improved specificity (76.72% vs. 70%) (57). In a similar context, Xu et al. developed a CNN-based model (KD-CNN) to distinguish KD from other pediatric illnesses. They analyzed 2,035 photographs of clinical signs (1,023 KD, 1,012 non-KD) from both public datasets and crowdsourced images. The model achieved an AUC of 0.90, with sensitivity of 0.80 and specificity of 0.85 (31).

Focusing on a different outcome, Lam et al. (2023) used CNNs and Vision Transformers to screen for KD. They analyzed 1,904 crowdsourced images of five clinical signs and validated on 605 images from 164 patients at Rady Children’s Hospital. Their models achieved 90% mean accuracy during cross-validation and 88% during external validation (32).

### Retinal Vasculitis Detection

Three studies focused on Retinal Vasculitis Detection. Amiot et al. analyzed 3,205 fluorescein angiography images from 148 patients (242 eyes). Their Pasa model, a CNN, achieved 90.5% accuracy in grading retinal vasculitis (59). Keino et al. used U-Net for automated detection of retinal vascular leakage, analyzing 12 ultrawide field fluorescein angiography images from 6 patients. They achieved a Dice coefficient of 0.467 for segmentation performance (70). Similarly, Dhirachaikulpanich et al. employed DeepLabV3+ and UNet++ models for segmenting retinal vascular leakage and occlusion. They analyzed 463 fluorescein angiography images from 82 patients, achieving Dice scores of 0.628 for leakage detection and 0.699 for occlusion detection (74).

### Other Vasculitis Imaging

Lee et al. (2022) used SE-ResNext50 to differentiate incomplete KD from pneumonia, analyzing 203 echocardiographic images (138 KD, 65 pneumonia). They achieved 75.86% accuracy, with sensitivity comparable to experienced cardiologists (57). Regarding other outcomes in KD, Luo et al. improved dual-source CT image segmentation for diagnosing coronary artery lesions in KD. They analyzed 90 children using a *RISEU-Net* model, achieving 96.7% accuracy in lesion segmentation (56). Lam et al. (2022) developed *KIDMATCH*, a two-stage deep learning model to differentiate multisystem inflammatory syndrome in children, KD, and other febrile illnesses. They analyzed data from 1,517 patients for internal validation and 175 for external validation. The model achieved an AUC of 98.8% in stage 1 (MIS-C identification) and 96.0% in stage 2 (KD identification), outperforming traditional clinician diagnosis (35).

### NLP

Three studies utilized NLP techniques to identify vasculitis cases from unstructured clinical text data.

Van Leeuwen et al. employed an NLP tool called *CTcue Patient Finder* to identify patients with ANCA-associated vasculitis in electronic health records, analyzing over 2 million records. The tool achieved a sensitivity of 97.0% in the training center and 97.6% in the validation center. The PPV improved significantly when using NLP, increasing from 56.9% to 77.9% in the training center and from 58.2% to 86.1% in the validation center. This NLP approach outperformed traditional ICD-10 coding methods (73).

Siewert et al. used the Dominance-Based Rough Set Approach (DRSA) for the differential diagnosis of KD. They analyzed data from 150 patients (48 with KD, 49 with infectious mononucleosis, and 53 with S. pyogenes pharyngitis), using demographic, physical examination, and laboratory data. The DRSA method generated 45 decision rules for diagnosing KD. The most accurate rule set achieved 100% sensitivity in recognizing KD without false negatives. However, the study did not report specificity or other performance metrics for these rule sets. The combination of thrombocytosis with ESR ≥ 77 mm/h or CRP ≥ 40.1 mg/L provided the highest diagnostic sensitivity for KD (42). Similarly, Doan et al. developed KD-NLP, a custom NLP tool to identify patients with high suspicion of KD from emergency department notes. They analyzed 253 patient records (166 from one site and 87 from another) based on the presence of fever and three or more clinical signs of KD. The tool achieved a sensitivity of 93.6% and specificity of 77.5%. In comparison, a simple keyword search method had a sensitivity of 41.0% and specificity of 76.3%. The KD-NLP tool performed comparably to manual chart review by physicians in identifying pediatric patients with a high suspicion of KD (41).

### Validation

Validation methods were reported in almost all of the studies (45/46). Internal validation techniques such as cross-validation and split datasets were more commonly used. External validation was performed in 10 studies.

For instance, Lam et al. (2022) validated KIDMATCH using external datasets from multiple hospitals, maintaining high accuracy across institutions (35). Van Leeuwen et al. conducted validation across two centers for the NLP model identifying ANCA-associated vasculitis (73). Pan et al. used a separate validation group to test the Random Forest model for renal damage prediction, achieving an accuracy of 84.2% (65).

Similarly, Linder et al. validated their machine learning models on an independent monocenter cohort (55). Doan et al. (2016) evaluated the KD-NLP tool against manually reviewed notes by physicians across two clinical sites (41).

## Discussion

This systematic review aimed to evaluate the current applications of AI in vasculitis, focusing on diagnosis and prediction tasks. We included 46 studies employing various AI methods across diverse data types. The findings revealed promising diagnostic performance for AI models, particularly in diagnosing KD, differentiating between vasculitis subtypes, and predicting treatment resistance and disease complications.

The results show that AI models have demonstrated strong potential use in specific cases of vasculitis diagnostics, particularly in diagnosing KD and predicting treatment outcomes. For instance, models using clinical and laboratory data achieved sensitivity as high as 92.5% and specificity up to 97.3% in distinguishing KD from other febrile conditions, suggesting that AI could be integrated into clinical workflows for early KD diagnosis. Predictive models for complications like IVIG resistance in KD have also performed well, with AUCs ranging from 0.716 to 0.834, highlighting their potential for identifying high-risk patients who may need closer monitoring or alternative therapies. These models could be integrated clinically by providing decision support for clinicians, enhancing patient stratification based on risk. Identifying patients at high risk of IVIG resistance is important because early detection allows for timely intervention with second-line treatments, potentially reducing the risk of coronary artery complications and improving long-term outcomes.

However, other applications of AI, such as imaging-based models, were less successful outside of KD. Although these models sometimes achieved high accuracy (e.g., 96% in coronary artery lesion detection for KD), their utility across other vasculitis types remains limited. One potential reason is the scarcity of large, diverse datasets for training AI models in rare vasculitis subtypes. The variability in imaging data, coupled with the rarity of these diseases, may hinder the models’ generalizability and their clinical adoption.

In contrast, AI models applied to broader vasculitis types, like predicting renal damage in IgA vasculitis, showed promising results, with AUCs reaching 0.91, but these were often based on small datasets, limiting their clinical readiness. The disparity in success between structured clinical data and imaging applications underscores the importance of expanding dataset sizes and diversity for conditions beyond KD. Furthermore, these results indicate that structured data models are closer to clinical integration, while imaging and other applications require further refinement.

Comparing these results with AI applications in other diseases, AI models for vasculitis remain less developed. In contrast to fields like oncology, cardiovascular disease, and broader rheumatologic conditions, where LLMs like GPT, BERT, and Claude have been more extensively researched (18,76,77), vasculitis AI applications have not yet explored these advanced models. LLMs offer promise not only in analyzing large volumes of unstructured clinical data but also, particularly with generative AI, in generating structured, human-like outputs that can be useful for healthcare professionals and patients. This capability could support earlier diagnosis and improve treatment personalization. Given some of the encouraging results of ML and DL models in vasculitis diagnostics, incorporating more recent models like LLMs could further improve diagnostic accuracy.

Despite these promising findings, this review reveals several limitations within the systematic literature review itself. First, the exclusion of non-English papers and preprints may have led to the omission of relevant studies, especially in such a fast-paced field. Additionally, the heterogeneity between studies—across AI models, datasets, and performance metrics—hindered our ability to conduct a meta-analysis.

Additionally, there are several limitations in the broader application of AI in vasculitis care, as highlighted by the current evidence. Many studies relied on small, single-center datasets, which limits the generalizability of their findings. Furthermore, external validation was conducted in only 10 studies, raising concerns about the robustness of these models when applied to broader clinical populations. The absence of newer AI models, such as LLMs, indicates a gap in the field that should be addressed to enhance AI’s clinical utility in vasculitis.

In conclusion, AI shows promising potential in diagnosing and predicting outcomes in vasculitis, offering the possibility of earlier diagnosis and intervention in these challenging, rare diseases. However, the current evidence is limited by small datasets, a lack of external validation, and the absence of advanced AI techniques such as LLMs. Future research should prioritize larger, multi-center studies, integrate newer AI models, and conduct extensive external validation to ensure clinical applicability.

## Data Availability

All data produced in the present study are available upon reasonable request to the authors

## Acknowledgment

none.

## Competing interest

None declared.

**Ethical approval** was not required for this research.

## Data Sharing Statement

This manuscript is a systematic review; therefore, all data utilized are available in the published articles that were reviewed. As this study did not generate new data, but rather analyzed existing published data, there are no additional datasets to share.

## Booleans used in each database

### PubMed

(((“artificial intelligence”[Title/Abstract] OR “AI”[Title/Abstract] OR “machine learning”[Title/Abstract] OR “ML”[Title/Abstract] OR “deep learning”[Title/Abstract] OR “neural networks”[Title/Abstract] OR “natural language processing”[Title/Abstract] OR “NLP”[Title/Abstract] OR “large language models”[Title/Abstract] OR “LLMs”[Title/Abstract] OR “computational modeling”[Title/Abstract] OR “supervised learning”[Title/Abstract] OR “unsupervised learning”[Title/Abstract] OR “reinforcement learning”[Title/Abstract])) AND ((“vasculitis”[Title/Abstract] OR “large vessel vasculitis”[Title/Abstract] OR “giant cell arteritis”[Title/Abstract] OR “Takayasu arteritis”[Title/Abstract] OR “medium vessel vasculitis”[Title/Abstract] OR “polyarteritis nodosa”[Title/Abstract] OR “Kawasaki disease”[Title/Abstract] OR “small vessel vasculitis”[Title/Abstract] OR “ANCA-associated vasculitis”[Title/Abstract] OR “granulomatosis with polyangiitis”[Title/Abstract] OR “microscopic polyangiitis”[Title/Abstract] OR “eosinophilic granulomatosis with polyangiitis”[Title/Abstract] OR “Henoch-Schönlein purpura”[Title/Abstract] OR “cryoglobulinemic vasculitis”[Title/Abstract] OR “cutaneous vasculitis”[Title/Abstract] OR “Behcet’s disease”[Title/Abstract] OR “central nervous system vasculitis”[Title/Abstract] OR “primary CNS vasculitis”[Title/Abstract] OR “secondary vasculitis”[Title/Abstract] OR “vasculitis syndrome”[Title/Abstract] OR “immune-mediated vasculitis”[Title/Abstract]))) AND (“research”[Title/Abstract] OR “study”[Title/Abstract] OR “trial”[Title/Abstract]) NOT (“review”[Publication Type] OR “comment”[Publication Type] OR “editorial”[Publication Type] OR “letter”[Publication Type])

### Embase

#2 (((“artificial intelligence”:ti,ab OR AI:ti,ab OR “machine learning”:ti,ab OR ML:ti,ab OR “deep learning”:ti,ab OR “neural networks”:ti,ab OR “natural language processing”:ti,ab OR NLP:ti,ab OR “large language models”:ti,ab OR LLMs:ti,ab OR “computational modeling”:ti,ab OR “supervised learning”:ti,ab OR “unsupervised learning”:ti,ab OR “reinforcement learning”:ti,ab))

`AND`

((“vasculitis”:ti,ab OR “large vessel vasculitis”:ti,ab OR “giant cell arteritis”:ti,ab OR “Takayasu arteritis”:ti,ab OR “medium vessel vasculitis”:ti,ab OR “polyarteritis nodosa”:ti,ab OR “Kawasaki disease”:ti,ab OR “small vessel vasculitis”:ti,ab OR “ANCA-associated vasculitis”:ti,ab OR “granulomatosis with polyangiitis”:ti,ab OR “microscopic polyangiitis”:ti,ab OR “eosinophilic granulomatosis with polyangiitis”:ti,ab OR “Henoch-Schonlein purpura”:ti,ab OR “cryoglobulinemic vasculitis”:ti,ab OR “cutaneous vasculitis”:ti,ab OR “Behcet disease”:ti,ab OR “central nervous system vasculitis”:ti,ab OR “primary CNS vasculitis”:ti,ab OR “secondary vasculitis”:ti,ab OR “vasculitis syndrome”:ti,ab OR “immune-mediated vasculitis”:ti,ab)))

`AND`

`([article]/lim OR [clinical trial]/lim OR [original research]/lim)`

`NOT ([editorial]/lim OR [letter]/lim OR [note]/lim OR [review]/lim)`

#2 AND (2000:py OR 2001:py OR 2002:py OR 2003:py OR 2004:py OR 2005:py OR 2006:py OR 2007:py OR 2008:py OR 2009:py OR 2010:py OR 2011:py OR 2012:py OR 2013:py OR 2014:py OR 2015:py OR 2016:py OR 2017:py OR 2018:py OR 2019:py OR 2020:py OR 2021:py OR 2022:py OR 2023:py OR 2024:py) AND [embase]/lim NOT ([embase]/lim AND [medline]/lim) AND (’article’/it OR ‘conference paper’/it OR ‘preprint’/it)

### Web of science

(TS=(“artificial intelligence” OR “machine learning” OR “deep learning” OR “neural networks” OR “natural language processing” OR NLP OR “large language models” OR LLMs OR GPT OR “GPT-3” OR “GPT-3.5” OR “GPT-4” OR “GPT-4o” OR Claude OR “computational modeling” OR “supervised learning” OR “unsupervised learning” OR “reinforcement learning”)

`AND`

TS=(“vasculitis” OR “large vessel vasculitis” OR “giant cell arteritis” OR “Takayasu arteritis” OR “medium vessel vasculitis” OR “polyarteritis nodosa” OR “Kawasaki disease” OR “small vessel vasculitis” OR “ANCA-associated vasculitis” OR “granulomatosis with polyangiitis” OR “microscopic polyangiitis” OR “eosinophilic granulomatosis with polyangiitis” OR “Henoch-Schonlein purpura” OR “cryoglobulinemic vasculitis” OR “cutaneous vasculitis” OR “Behcet disease” OR “central nervous system vasculitis” OR “primary CNS vasculitis” OR “secondary vasculitis” OR “vasculitis syndrome” OR “immune-mediated vasculitis”))

### Scopus

((TITLE (“artificial intelligence” OR ai OR “machine learning” OR ml OR “deep learning” OR “neural networks” OR “natural language processing” OR nlp OR “large language models” OR llms OR gpt OR “GPT-3” OR “GPT-3.5” OR “GPT-4” OR “GPT-4o” OR claude OR “computational modeling” OR “supervised learning” OR “unsupervised learning” OR “reinforcement learning”)) AND (TITLE (“vasculitis” OR “large vessel vasculitis” OR “giant cell arteritis” OR “Takayasu arteritis” OR “medium vessel vasculitis” OR “polyarteritis nodosa” OR “Kawasaki disease” OR “small vessel vasculitis” OR “ANCA-associated vasculitis” OR “granulomatosis with polyangiitis” OR “microscopic polyangiitis” OR “eosinophilic granulomatosis with polyangiitis” OR “Henoch-Schonlein purpura” OR “cryoglobulinemic vasculitis” OR “cutaneous vasculitis” OR “Behcet disease” OR “central nervous system vasculitis” OR “primary CNS vasculitis” OR “secondary vasculitis” OR “vasculitis syndrome” OR “immune-mediated vasculitis”))) AND (LIMIT-TO (DOCTYPE, “ar”))

### IEEE Xplore

((“artificial intelligence” OR “machine learning” OR “deep learning” OR “neural networks” OR “natural language processing” OR NLP OR “large language models” OR LLMs OR GPT OR “GPT-3” OR “GPT-3.5” OR “GPT-4” OR “GPT-4o” OR Claude OR “computational modeling” OR “supervised learning” OR “unsupervised learning” OR “reinforcement learning”)

`AND`

(“vasculitis” OR “large vessel vasculitis” OR “giant cell arteritis” OR “Takayasu arteritis” OR “medium vessel vasculitis” OR “polyarteritis nodosa” OR “Kawasaki disease” OR “small vessel vasculitis” OR “ANCA-associated vasculitis” OR “granulomatosis with polyangiitis” OR “microscopic polyangiitis” OR “eosinophilic granulomatosis with polyangiitis” OR “Henoch-Schonlein purpura” OR “cryoglobulinemic vasculitis” OR “cutaneous vasculitis” OR “Behcet disease” OR “central nervous system vasculitis” OR “primary CNS vasculitis” OR “secondary vasculitis” OR “vasculitis syndrome” OR “immune-mediated vasculitis”))

