## Supplementary material for "Applications of Artificial Intelligence in Vasculitides: A Systematic Review": Tables 1-2

**Table 1**: A summary of the included studies.

| Author | Year | AI Method | Type of Vasculitis | Task | Specific Task | Model Performance |
|---|---|---|---|---|---|---|
| **Vries et al.** | 2023 | ML (XGBoost) | GCA | Diagnosis | Differentiate GCA from atherosclerosis | AUC-ROC: 0.96, Accuracy: 90% |
| **Ryyppö et al.** | 2024 | ML (XGBoost) | Various | Diagnosis | Early identification of vasculitides | AUC: 0.808, TPR: 76.7%, TNR: 98.4% |
| **Chi Li et al.** | 2023 | ML (LASSO, SVM) | KD | Diagnosis | Distinguish KD from sepsis | AUC: 0.926 (training), 0.878 (validation) |
| **Zhang et al.** | 2022 | ML (RF, ANN) | KD | Diagnosis | Distinguish KD from healthy/convalescent | AUC: 1.0 (KD), 0.945 (convalescent) |
| **Astion et al.** | 1994 | ML (NN) | GCA | Diagnosis | Classify GCA vs other vasculitis | Accuracy: 94.4% (GCA), 91.9% (other) |
| **Hammam et al.** | 2023 | ML (XGBoost) | Behçet's | Prediction | Predict vision-threatening BD | AUC: 0.85, Accuracy: 85% |
| **Tsai et al.** | 2023 | ML (XGBoost) | KD | Diagnosis | Differentiate KD from febrile children | AUC: 0.980, Sensitivity: 92.5%, Specificity: 97.3% |
| **Wang et al.** | 2024 | ML (RF, SVM) | KD | Prediction | Predict IVIG resistance | AUC: 0.816 (training), 0.800 (validation) |
| **Sunaga et al.** | 2023 | ML (LightGBM) | KD | Prediction | Predict IVIG resistance | AUC: 0.78, Sensitivity: 0.50, Specificity: 0.88 |
| **Cao et al.** | 2022 | ML (XGBoost) | HSP | Prediction | Predict renal damage | AUC: 0.895 (training), 0.870 (test) |
| **Lu et al.** | 2023 | ML (bagFDAGCV) | Takayasu | Prediction | Predict ischemic complications | AUC: 0.773 |
| **Ing et al.** | 2019 | ML (NN, LR) | GCA | Prediction | Predict biopsy-proven GCA | AUC: 0.860 (NN), Sensitivity: 69.5%, Specificity: 89.1% |

| | | | | | | |
|---|---|---|---|---|---|---|
| **Venerito et al.** | 2022 | ML (RF) | GCA | Prediction | Predict GCA flare | AUC: 0.76, Accuracy: 71.4% |
| **Tang et al.** | 2024 | ML (RF) | KD | Prediction | Predict coronary artery lesions | AUC: 0.925, Accuracy: 93% |
| **Joung et al.** | 2022 | ML (Decision Tree) | KD | Prediction | Predict IVIG resistance and CAD | Accuracy: 90.5% (IVIG), 90.3% (CAD) |
| **Duan et al.** | 2024 | ML (EBM) | KD | Diagnosis | Diagnose KD | AUC: 0.97, Accuracy: 93% |
| **Nie et al.** | 2023 | ML (XGBoost) | HSP | Diagnosis | Differentiate AHSP from acute appendicitis | AUC: 0.895, Accuracy: 82.4% |
| **Güler et al.** | 2005 | ML (MLPNN) | Behçet's | Diagnosis | Detect ocular Behçet's disease | Accuracy: 96.43% (healthy), 93.75% (Behçet's) |
| **Wang et al.** | 2022 | ML (RF) | IgA Vasculitis | Prediction | Predict renal damage | Accuracy: 0.83, AUC: 0.91 |
| **Miyagi et al.** | 2024 | ML (Stacking) | KD | Prediction | Predict IVIG non-responsiveness | AUC: 0.716, Sensitivity: 0.691, Specificity: 0.629 |
| **Azuma et al.** | 2020 | ML (NN) | KD | Prediction | Predict coronary artery lesions | Sensitivity: 73%, Specificity: 99%, C-index: 0.86 |
| **Pan et al.** | 2024 | ML (RF) | IgA Vasculitis | Prediction | Predict renal damage | Accuracy: 91%, AUC: 0.94 |
| **Morris et al.** | 2022 | ML (PCA-LDA, SVM) | AAV | Diagnosis | Detect histological lesions | ROC-AUC: 0.97-1.00 |
| **Liu et al.** | 2021 | ML (LightGBM) | KD | Prediction | Predict IVIG resistance | AUC: 0.874, Sensitivity: 0.702, Specificity: 0.903 |
| **Kuniyoshi et al.** | 2020 | ML (LR, SVM, XGB) | KD | Prediction | Predict IVIG resistance | AUC: 0.58-0.75 |
| **Deng et al.** | 2024 | ML (XGBoost) | KD | Prediction | Predict IVIG resistance | AUC: 0.821, Accuracy: 0.748 |
| **Lu et al.** | 2024 | ML (NN) | Behçet's | Prediction | Predict Behçet's disease uveitis | AUC: 0.900, Accuracy: 76.5% |

| | | | | | | |
|---|---|---|---|---|---|---|
| **Takeuchi et al.** | 2017 | ML (RF) | KD | Prediction | Predict IVIG resistance | AUC: 0.916, Sensitivity: 79.7%, Specificity: 87.3% |
| **Schmitt et al.** | 2001 | ML (ANN) | EGPA, GPA | Diagnosis | Differentiate CSS from WG | Accuracy: 96% (validation) |
| **Ren et al.** | 2024 | ML (RF) | IgA Nephropathy | Diagnosis | Classify IgAN subtypes | Accuracy: 96.15%, AUC: 0.8775 (average) |
| **Abdolmanafi et al.** | 2017 | DL (CNN) | KD | Diagnosis | Classify coronary artery layers | Accuracy: 96-97% |
| **Lam et al.** | 2023 | DL (CNN, ViT) | KD | Diagnosis | Screen KD using clinical signs | Accuracy: 90% (cross-validation), 88% (external) |
| **Amiott et al.** | 2023 | DL (CNN) | Retinal vasculitis | Diagnosis | Grade retinal vasculitis | F1 score: 0.81, AUC: 0.86 |
| **Lee et al.** | 2024 | DL (MRANet) | KD | Diagnosis | Differentiate incomplete KD from pneumonia | Accuracy: 78.82% |
| **Rodríguez et al.** | 2022 | DL (U-Net) | KD | Diagnosis | Segment coronary arteries | Accuracy: 99.76%, F1-score: 68.08 |
| **Xu et al.** | 2022 | DL (CNN) | KD | Diagnosis | Differentiate KD from other illnesses | AUC: 0.90, Sensitivity: 0.80, Specificity: 0.85 |
| **Keino et al.** | 2022 | DL (U-Net) | Retinal vasculitis | Diagnosis | Detect retinal vascular leakage | Precision: 0.434, Recall: 0.529, Dice: 0.467 |
| **Dhirachaikulpanich et al.** | 2024 | DL (UNet++) | Retinal vasculitis | Diagnosis | Segment retinal vascular leakage | Dice score: 0.6279 (leakage), 0.6992 (occlusion) |
| **Lee et al.** | 2022 | DL (SE-ResNext50) | KD | Diagnosis | Distinguish incomplete KD from pneumonia | Accuracy: 75.86%, AUC: 0.8563 |
| **Luo et al.** | 2022 | DL (CNN) | KD | Diagnosis | Segment coronary artery lesions | Accuracy: 96.7%, Sensitivity: 95.6% |

| | | | | | | |
|---|---|---|---|---|---|---|
| **Lam et al.** | 2022 | DL (NN) | KD, MIS-C | Diagnosis | Differentiate KD, MIS-C, and other illnesses | AUC: 98.8% (MIS-C), 96.0% (KD) |
| **Linder et al.** | 2011 | DL (ANN) | GPA, MPA | Diagnosis | Differentiate WG from MPA | Accuracy: 94.3% (multicenter), 91% (monocenter) |
| **Abdolmanafi et al.** | 2020 | DL (CNN, FCN) | KD | Diagnosis | Diagnose coronary artery lesions | Accuracy: 90-95% |
| **van Leeuwen et al.** | 2024 | NLP | AAV | Diagnosis | Identify AAV patients in EHRs | Sensitivity: 97.0%, PPV: 77.9-86.1% |
| **Siewert et al.** | 2021 | NLP (DRSA) | KD | Diagnosis | Differential diagnosis of KD | Sensitivity: 100% for specific rule sets |
| **Doan et al.** | 2016 | NLP | KD | Diagnosis | Identify KD suspicion in ED notes | Sensitivity: 93.6%, Specificity: 77.5% |

**Abbreviations**: ML: Machine Learning, DL: Deep Learning, NLP: Natural Language Processing GCA: Giant Cell Arteritis, KD: Kawasaki Disease, HSP: Henoch-Schönlein Purpura, AAV: ANCA-Associated Vasculitis, EGPA: Eosinophilic granulomatosis with polyangiitis, GPA: Granulomatosis with polyangiitis, MPA: Microscopic Polyangiitis AUC: Area Under the Curve, ROC: Receiver Operating Characteristic, TPR: True Positive Rate, TNR: True Negative Rate IVIG: Intravenous Immunoglobulin, CAD: Coronary Artery Dilatation, ED: Emergency Department NN: Neural Network, RF: Random Forest, SVM: Support Vector Machine, CNN: Convolutional Neural Network, ViT: Vision Transformer, FCN: Fully Convolutional Network MLPNN: Multilayer Perceptron Neural Network, EBM: Explainable Boosting Machine, DRSA: Dominance-Based Rough Set Approach

**Table 2**: Summary of the data types, metrics, limitations and implications of the included studies.

| Author | Year | Data Type | Sample Size | Performance Summary | Reported Limitations | Implications |
|---|---|---|---|---|---|---|
| **Vries et al.** | 2023 | Imaging data (FDG-PET) | 20 patients | AUC-ROC: 0.96, Accuracy: 90% | Small sample size, manual delineation variability | Improved differentiation between GCA and atherosclerosis |
| **Ryyppö et al.** | 2024 | Clinical data, lab results | 114,897 patients | AUC: 0.808, TPR: 76.7%, TNR: 98.4% | Some patients had pre-existing diagnoses | Early identification of vasculitides, reducing diagnostic delay |
| **Chi Li et al.** | 2023 | Clinical data, lab results | 608 patients | AUC: 0.926 (training), 0.878 (validation) | Single-center study, retrospective design | Improved differentiation between KD and sepsis |
| **Zhang et al.** | 2022 | Gene expression profiles | 474 samples across multiple datasets | AUC: 1.0 (KD), 0.945 (convalescent) | Clinical data heterogeneity across datasets | Effective distinction of KD from healthy and convalescent states |
| **Astion et al.** | 1994 | Clinical data | 807 cases | Accuracy: 94.4% (GCA), 91.9% (other) | Potential for overtraining | Improved diagnostic classification in complex diseases like GCA |
| **Hammam et al.** | 2023 | Clinical data | 1094 BD patients | AUC: 0.85, Accuracy: 85% | Cross-sectional design, potential collinearity | Early intervention for vision-threatening BD |
| **Tsai et al.** | 2023 | Clinical data | 74,641 patients | AUC: 0.980, Sensitivity: 92.5%, Specificity: 97.3% | Limited to Taiwanese population under 5 years | Early KD diagnosis in emergency departments |
| **Wang et al.** | 2024 | Clinical data, lab results | 1271 patients | AUC: 0.816 (training), 0.800 (validation) | Single-center study, small IVIG-resistant group | Early prediction of IVIG resistance in KD |
| **Sunaga et al.** | 2023 | Clinical data, lab results | 1002 KD cases | AUC: 0.78, Sensitivity: 0.50, Specificity: 0.88 | Primarily Japanese patients, retrospective data | Practical tool for predicting IVIG resistance in KD |
| **Cao et al.** | 2022 | Clinical data, lab results | 240 children with HSP | AUC: 0.895 (training), 0.870 (test) | Single-center study, small sample size | Non-invasive prediction of renal damage in HSP |
| **Lu et al.** | 2023 | Clinical data | 703 patients | AUC: 0.773 | Single-center design, lack of temporal data | Identification of high-risk Takayasu arteritis patients |
| **Ing et al.** | 2019 | Clinical data, lab results | 1,201 patients | AUC: 0.860 (NN), Sensitivity: 69.5%, Specificity: 89.1% | Missing data for bloodwork in 34% of cases | Improved triage for GCA biopsy |

| | | | | | | |
|---|---|---|---|---|---|---|
| **Venerito et al.** | 2022 | Clinical data, lab results | 107 GCA patients | AUC: 0.76, Accuracy: 71.4% | Small sample size, retrospective design | Prediction of GCA flare after glucocorticoid tapering |
| **Tang et al.** | 2024 | Clinical data | 158 children | AUC: 0.925, Accuracy: 93% | Lack of external validation, regional dataset | Early risk stratification for CAL in KD |
| **Joung et al.** | 2022 | Clinical data, lab results | 896 children | Accuracy: 90.5% (IVIG), 90.3% (CAD) | Limited to two centers | Early identification of high-risk KD patients |
| **Duan et al.** | 2024 | Clinical data | 4,087 pediatric patients | AUC: 0.97, Accuracy: 93% | Single hospital data, no external validation | Interpretable KD diagnosis model |
| **Nie et al.** | 2023 | Laboratory data | 6,965 patients | AUC: 0.895, Accuracy: 82.4% | No imaging data included | Differentiation between AHSP and acute appendicitis |
| **Güler et al.** | 2005 | Ophthalmic arterial Doppler signals | 106 subjects | Accuracy: 96.43% (healthy), 93.75% (Behçet's) | Limited to ophthalmic signals | Rapid detection of ocular Behçet's disease |
| **Wang et al.** | 2022 | Clinical data | 288 children | Accuracy: 0.83, AUC: 0.91 | Single-center study, limited sample size | Early prediction of renal damage in IgA vasculitis |
| **Miyagi et al.** | 2024 | Clinical data, lab results | 1225 patients | AUC: 0.716, Sensitivity: 0.691, Specificity: 0.629 | Retrospective design, data quality issues | Prediction of IVIG non-responsiveness in KD |
| **Azuma et al.** | 2020 | Clinical data | 314 KD patients | Sensitivity: 73%, Specificity: 99%, C-index: 0.86 | Small validation sample size | Prediction of CAL development in KD |
| **Pan et al.** | 2024 | Clinical data | 263 patients | Accuracy: 91%, AUC: 0.94 | Single-center design, small sample size | Early identification of renal damage in IgA vasculitis |
| **Morris et al.** | 2022 | Tissue and urine samples | 27 tissue samples, 10 urine samples | ROC-AUC: 0.97-1.00 | Small sample size, limited urine sample performance | Non-invasive monitoring of ANCA-associated glomerulonephritis |
| **Liu et al.** | 2021 | Clinical data, lab results | 1,398 patients | AUC: 0.874, Sensitivity: 0.702, Specificity: 0.903 | Retrospective analysis, potential missing data bias | Early identification of IVIG-resistant KD |
| **Kuniyoshi et al.** | 2020 | Clinical data, lab results | 98 children | AUC: 0.58-0.75 | Small dataset size, single-center data | Initial step in predicting IVIG resistance in KD |
| **Deng et al.** | 2024 | Clinical data | 602 patients | AUC: 0.821, Accuracy: 0.748 | Small sample size for validation, selection bias | Reliable tool for predicting IVIG-resistance in KD |
| **Lu et al.** | 2024 | Imaging data (OCTA scans), Clinical data | 123 patients | AUC: 0.900, Accuracy: 76.5% | Limited sample size, single population study | Early detection of Behçet's disease uveitis |

| | | | | | | |
|---|---|---|---|---|---|---|
| **Takeuchi et al.** | 2017 | Clinical data | 767 patients | AUC: 0.916, Sensitivity: 79.7%, Specificity: 87.3% | Lack of external validation, accessibility issues | Risk-tailored therapy for IVIG resistance in KD |
| **Schmitt et al.** | 2001 | Clinical data, lab results | 80 patients | Accuracy: 96% (validation) | Limited to two vasculitis types, no histology data | Improved differentiation between EGPA and GPA |
| **Ren et al.** | 2024 | Gene expression data | 107 IgAN patients, 14 controls | Accuracy: 96.15%, AUC: 0.8775 (average) | Lack of follow-up data | Personalized treatment guidance for IgAN |
| **Abdolmanafi et al.** | 2017 | OCT images | 26 patients, 4800 ROIs | Accuracy: 96-97% | Small dataset, overfitting concerns | Improved analysis of coronary abnormalities in KD |
| **Lam et al.** | 2023 | Clinical images | 2,509 images | Accuracy: 90% (cross-validation), 88% (external) | Limited external validation dataset | Early KD screening tool |
| **Amiott et al.** | 2023 | Fluorescein angiography images | 3,205 images, 148 patients | F1 score: 0.81, AUC: 0.86 | High variability in image intensity | Automated grading of retinal vasculitis |
| **Lee et al.** | 2024 | Echocardiographic images | 147 images | Accuracy: 78.82% | Limited dataset size, class imbalance | Assistive tool for incomplete KD diagnosis |
| **Rodríguez et al.** | 2022 | Echocardiography images | 1,531 images | Accuracy: 99.76%, F1-score: 68.08 | Small dataset, need for data augmentation | Automated coronary artery segmentation in KD |
| **Xu et al.** | 2022 | Clinical images | 2,035 images | AUC: 0.90, Sensitivity: 0.80, Specificity: 0.85 | Limited dataset, lack of demographic diversity | Aid in distinguishing KD from other illnesses |
| **Keino et al.** | 2022 | Fluorescein angiography images | 12 images from 6 patients | Precision: 0.434, Recall: 0.529, Dice: 0.467 | Small sample size, single institution data | Objective assessment of retinal vasculitis activity |
| **Dhirachaikulpanich et al.** | 2024 | Fluorescein angiography images | 463 images from 82 patients | Dice score: 0.6279 (leakage), 0.6992 (occlusion) | Retrospective nature, small dataset | Enhanced diagnostic precision in retinal vasculitis |
| **Lee et al.** | 2022 | Echocardiographic images | 203 images | Accuracy: 75.86%, AUC: 0.8563 | Small dataset, class imbalance | Assistive tool for incomplete KD diagnosis |
| **Luo et al.** | 2022 | CT images | 90 children | Accuracy: 96.7%, Sensitivity: 95.6% | Small sample size | Improved diagnosis of coronary artery lesions in KD |
| **Lam et al.** | 2022 | Clinical data | 1,517 patients (internal), 175 (external) | AUC: 98.8% (MIS-C), 96.0% (KD) | Reliance on pre-pandemic KD data | Clinical decision support for KD and MIS-C diagnosis |
| **Linder et al.** | 2011 | Clinical data | 318 patients (multicenter), 67 (monocenter) | Accuracy: 94.3% (multicenter), 91% (monocenter) | Limited to two vasculitis types | Improved differentiation between GPA and MPA |

| | | | | | | |
|---|---|---|---|---|---|---|
| **Abdolmanafi et al.** | 2020 | OCT images | 5040 frames from 45 pullbacks | Accuracy: 90-95% | Limited dataset, potential data imbalance | Improved diagnosis of coronary artery lesions in KD |
| **van Leeuwen et al.** | 2024 | EHR data | 2,000,000+ records | Sensitivity: 97.0%, PPV: 77.9-86.1% | Inability to calculate specificity and NPV | Improved identification of AAV patients in EHRs |
| **Siewert et al.** | 2021 | Clinical data, lab results | 150 patients | Sensitivity: 100% for specific rule sets | Small sample size, retrospective nature | Effective differential diagnosis of KD |
| **Doan et al.** | 2016 | ED notes | 253 patients | Sensitivity: 93.6%, Specificity: 77.5% | Limited syntactical structures in training data | Timely recognition of KD in emergency settings |

**Abbreviations**: ML: Machine Learning, DL: Deep Learning, NLP: Natural Language Processing GCA: Giant Cell Arteritis, KD: Kawasaki Disease, HSP: Henoch-Schönlein Purpura, AAV: ANCA-Associated Vasculitis, EGPA: Eosinophilic granulomatosis with polyangiitis, GPA: Granulomatosis with polyangiitis, MPA: Microscopic Polyangiitis, BD: Behçet's Disease, IgA: Immunoglobulin A, IgAN: IgA Nephropathy AUC: Area Under the Curve, ROC: Receiver Operating Characteristic, TPR: True Positive Rate, TNR: True Negative Rate, PPV: Positive Predictive Value, NPV: Negative Predictive Value IVIG: Intravenous Immunoglobulin, CAL: Coronary Artery Lesions, CAD: Coronary Artery Dilatation, ED: Emergency Department NN: Neural Network, RF: Random Forest, SVM: Support Vector Machine, CNN: Convolutional Neural Network, ViT: Vision Transformer, FCN: Fully Convolutional Network MLPNN: Multilayer Perceptron Neural Network, EBM: Explainable Boosting Machine, DRSA: Dominance-Based Rough Set Approach OCT: Optical Coherence Tomography, ROI: Region of Interest, CT: Computed Tomography, EHR: Electronic Health Record, MIS-C: Multisystem Inflammatory Syndrome in Children
